# Alzheimer’s disease polygenic risk associated dynamic functional networks and anatomical asymmetry

**DOI:** 10.1101/2023.07.05.23292258

**Authors:** Nicolas Rubido, Gernot Riedel, Vesna Vuksanović

**Affiliations:** Institute of Complex Systems and Mathematical Biology, University of Aberdeen, UK; Institute of Medical Sciences, University of Aberdeen, UK; Swansea University Medical School, Health Data Science, Data Science Building, Swansea, SA2 8PP, United Kingdom

**Keywords:** dynamic functional connectivity, polygenic risk, cholinergic pathways, Alzheimer’s disease

## Abstract

Genetic associations with macroscopic brain networks can provide insights into healthy and aberrant cortical connectivity in disease. However, associations specific to dynamic functional connectivity in Alzheimer’s disease are still largely unexplored. Understanding the association between gene expression in the brain and functional connectivity may provide useful information about the molecular processes underlying variations in impaired brain function. Given the potential of dynamic functional connectivity to uncover brain states associated with Alzheimer’s disease, it is interesting to ask: How does gene expression associated with Alzheimer’s disease map onto the dynamic functional brain connectivity? If genetic variants associated with neurodegenerative processes involved in Alzheimer’s disease are to be correlated with brain function, it is essential to generate such a map. Here, we investigate how the relation between gene expression in the brain and dynamic functional connectivity arises from nodal interactions, quantified by their role in network centrality (i.e., the drivers of the metastability), and the principal component of genetic co-expression across the brain. Our analyses include genetic variations associated with Alzheimer’s disease and also genetic variants expressed within the cholinergic brain pathways. Our findings show that contrasts in metastability of functional networks between Alzheimer’s and healthy individuals can in part be explained by the two combinations of genetic co-variations in the brain with the confidence interval between 72% and 92%. The highly central nodes, driving the brain aberrant metastable dynamics in Alzheimer’s disease highly correlate with the magnitude of variations of the two combinations of genes expressed in brain. These nodes include mainly the white matter, parietal and occipital brain regions, each of which (or their combinations) are involved in impaired cognitive function in Alzheimer’s disease. In addition, our results provide evidence of the role of genetic associations across brain regions in asymmetric changes in ageing. We validated our findings on the same cohort using alternative brain parcellation methods. This work demonstrates how genetic variations underpin anomaly in dynamic functional connectivity in Alzheimer’s disease.

## Introduction

Brain network analysis from resting-state functional MRI (fMRI) has advanced our understanding of the cortical organization in healthy brain (1). Recently, dynamic brain networks’ analysis, which considers temporal fluctuations in the resting-state fMRI signal, has revealed patterns of activity, that are usually averaged out by conventional functional network analysis. These patterns of activity, termed dynamic networks, reveal transient (meta-stable) dynamical states, likely involved in cognitive processing (2). In this context, dynamic functional networks analysis from fMRI data has shown a potential to unveil clinically relevant information (1, 3). Capturing the evolving architecture of brain networks over short periods of time might also provide pathophysiological insights into neurological and neurodegenerative conditions, and will provide better diagnostic or prognostic indicators.

The non-stationary nature of the brain organization and the differences across healthy and diseased cohorts, have been widely investigated by means of variations of the dwell time within different sub-network configurations (4). For example, a study that investigated the evolution of dynamical Functional Connectivity (dFC) disruptions across the AD spectrum, has shown differences in patients with dementia compared to Mild Cognitive Impairment (MCI) in terms of local dFC within the temporal, frontal-superior and default-mode sub-networks. Moreover, the decreased global metastability between functional states has also been reported (5, 6). Consistently, studies showed a progressive loss of whole-brain metastability according to the severity of cognitive impairment along the AD continuum, reaching statistical significance only in patients with dementia, when compared with healthy controls (7, 8). These findings support the hypothesis that global patterns of brain activity in AD are progressively altered, and eventually lead to a loss of ‘dynamic complexity’ (i.e., the decrease in possible functional networks configurations). However, neurobiological correlates of such differences have remained elusive.

Alzheimer’s disease is a complex, an irreversible neurodegenerative disorder, which causes cognitive impairments leading to dementia. Age is the greatest risk factor, but many other risk factors have been identified and associated with AD (9). Genome-wide association studies (GWAS) have identified numerous genetic variants with small cumulative effects, which have been aggregated into a polygenic risk score (PRS) that may explain up to 58–79% of AD heritability and earlyonset AD showing over 90% (10). In addition, it has been suggested that the significant polygenic component of AD risk, could be “a valuable research tool complementing experimental designs” (11). Any of these genes contribute to amyloid accumulation, tau protein misfolding, the innate immune response, regulation of endocytosis, and proteasome-ubiquitin activity (12–14). This pathology propagates along predefined neuronal pathways (15). Indeed, functional brain imaging (fMRI and PET) and spatial patterns of neurodegeneration in AD mirror the anatomy of functional brain networks (16–19), of which the Braak staging is the most prominent candidate (20). Based on the tau protein aggregation pattern along interconnected neuronal pathways, they have defined 6 severity stages. Stages >3 present with the onset of AD-like symptoms and correlate with pathology in entorhinal and hippocampal areas, from which further cortical spread leads to increased deterioration of the patients. We therefore here explored whether any genetic associations related to AD are explainable by functional brain networks analysis and, in particular, their non-stationary nature. This selective pattern of neurodegeneration can be seen in anatomical MRI studies from AD patients (16, 21), which have confirmed the Braak staging with the onset of tissue loss in the entorhinal area, the hippocampus, the ventral striatum and the basal part of the forebrain in early stages of AD (22). Especially the latter brain structures are well known for their high content of cholinergic neurones (23) with long-reaching afferents terminating in the cortical mantle and hippocampus. These projections play a critical role in learning and memory processes (24, 25) and are severely diminished in patients with AD (26, 27), which prompted the development of cholinergic treatments as therapy for AD patients. The two views are not mutually exclusive as tau also aggregates in cholinergic cells (28) and presumably terminals (29) thereby compromising cholinergic tone in these target structures. Since these also comprise cortical elements their inclusion in and correlation with the dFC network metrics appears warranted.

## Methods

### A. Participants and Cohorts

Data used in this study were obtained from the Alzheimer’s Disease Neuroimaging Initiative [(ADNI)](https://adni.loni.usc.edu/) data base. We downloaded demographic, clinical and MRI data from *N* = 315 participants. The main inclusion criteria was that the fMRI datasets were acquired using the same resting-state protocols (see the Imaging Datasets Section). The 315 participants were selected from Healthy Controls (HC), Mild cognitive Impairment (MCI), and Alzheimer’s Disease (AD) groups. Demographic and clinical characteristics of the participant averaged across each group were shown in Table 1.

**Table 1.** Participants demographic and clinical characteristics.

| Demographics | Healthy | MCI | AD |
| --- | --- | --- | --- |
| Number | 141 | 128 | 46 |
| Sex (F/M) | 68/73 | 52/76 | 17/29 |
| Age (sd) – years | 80(6) | 77(5)* | 80(6) |
| Clinical Scores |  |  |  |
| MMSE | 28(4) | 27(3) | 22(4)*,δ |

Abbreviations: AD – Alzheimer's Disease, MCI – Mild Cognitive Impairment; F – Females; M – Males; sd – standard deviation. \* significant difference (i.e., $p < 0.01$ ) when comparing with HC. δ significant difference (i.e., $p < 0.01$ ) when comparing with MCI.

#### Imaging Datasets and Processing

We analysed restingstate functional MRI (rs-fMRI, or fMRI in the text) data from 315 individuals. fMRI were acquired using the ADNI-3 basic EPI-BOLD protocol (details at [ADNI Imaging Protocol](http://adni.loni.usc.edu/data-samples/data-types/mri)) and in (30). In short, the participants had their scans taken for up to 10 minutes using the same two-time accelerated 3 *T* scanners, following an even/odd interleaved axial-slicing (inferior to superior) of 3.4 *mm* with (3.4375 *mm*)2 pixels (FOV = 220 × 220 × 163*mm*; *P >> A* phase encoding; TE = 30 *ms*; TR = 3000 *ms*).

Image pre-processing, including brain extraction, registration to standard MNI space, and brain tissue segmentation was carried using FMRIB’s pipeline fsl_anat (31) at its default settings. Pre-processing of fMRI was done by applying the FMRIB’s Expert Analysis Tool, FEAT, resulting in voxel-wise Blood-Oxygen Level-Dependent (BOLD) signals of *N*_*T*_ = 197 data points, or in 197×TR acquisition times.

#### Construction of Dynamic Functional Networks

Cortical regions – defining the network nodes – are based on either the Juelich (JBA) (32) or the Harvard-Oxford (H-OBA) (33) Brain Atlas for each participant’s fMRI. This resulted in 2 cortical parcellations of *N*_*node*_ = (48 + 21) or 121 nodes (regions/parcels), respectively. Details of the atlases in Table 5. JBA is a three-dimensional atlas containing cytoarchitectonic maps of cortical areas and subcortical nuclei. The atlas is probabilistic, which enables it to account for variations between individual brains. The signal of each node is determined by averaging fMRI BOLD signals across the regional voxels. We then define a functional link by the Pearson’s correlation coefficient, *ρ*_*i,j*_, between all possible node pairs (node - *i*; and node - *j*). See for example our earlier work (2), for details on networks construction from restingstate fMRI data. We also set a 99% significance threshold to the value of each pair-wise correlation to remove spurious correlations. That is, all pair-wise correlations that were not significant at *p <* 0.01 were excluded from any further analyses. This definition of functional links as a pair-wise correlation between regional BOLD time-series, results in a single-subject, symmetric (undirected), weighted (with positive and negative weights), functional network that was build on statistically significant (temporal) interactions between its nodes.

In more details, the dynamic functional network (dFN) of each participant, was built using half-overlapping sliding windows of the node signals. Specifically, *t*_*m*_ = *m*Δ*t/*2, with *m* = 0, 1,… *< N*_*T*_ */*Δ*t*, and Δ*t* = 20 data points – accounting to 1 minutes of scan time. The exploration of window length on dFC networks and their characteristics, as well as optimal window-length for the analysis of fMRI data has been described previously (34). Here, *t*_*m*_ is a sliding time-window, whose (sliding) properties were defined by a multiplication factor (*m*) of the window length (Δ*t*), and the full length of the BOLD signal (*N*_*T*_). As a result, *ρ*_*i,j*_ changes in time depending on the start *t*_*m*=0_ of the sliding window. We use the significance threshold from the FC on the entire BOLD time series, given by *ρ*_*i,j*_(*t*_*T*_) for each participant, to threshold the dFN *ρ*_*i,j*_(*t*_*m*_). This corresponds to the sliding-window correlations being higher or equal than the corresponding correlations on the window from the entire BOLD signal (i.e. *t*_*T*_ = 197 * *TR*). Consequently, when using the H-OBA and a sliding window of Δ*t* = 20 points (resulting in 18 windows over the total signal), the average percentage of links discarded was 28%, whereas when using the JBA, the average percentage of discarded links per participant was 30%.

#### Characterisation of Functional Networks

We characterised functional connectivity in three study groups depending on the importance of the network nodes. A straightforward method of assessing the importance of a network node is to compute its centrality. The centrality of a node is a measure that quantifies how important or influential a node is within a network. Centrality can be expressed in various ways; thus there are multiple types of centrality measures. We characterised the importance of each node in the dynamic functional networks of HC, MCI, and AD individuals by calculating the node strength and eigenvector centrality. We analyse these two, basic centrality network measures from the *N*_*nodes*_ × *N*_*nodes*_ correlation matrices of each participant, based on using the sliding window approach described in the section above.

##### A.1 Node Strength

The simplest measure of network centrality is the so called degree centrality, or simply node degree, which represents the number of connections of a node to other nodes in the network. In weighted networks, this corresponds to the node strength, which is a measure of the strength of the functional connections of a node *i* (for a single *N*_*nodes*_ × *N*_*nodes*_ network). For dFC networks, the node strength is calculated using the equation below:

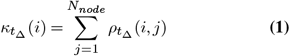

Where *i* and *j* are node indices, *t*_Δ_ = *t* ∈ [*t*_0_, *t*_0_ + Δ*t*] sliding time window of the size Δ*t*. 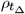 is an element of the correlation matrix representing the strength of the pair-wise correlation between pairs of nodes (*i, j*) within a sliding window *t*_Δ_.

##### A.2 Eigenvector Centralit

Eigenvector centrality is the centrality measure which is based on the assumption that connections to more influential nodes are more important than connections to less relevant nodes, the measure also takes the centrality of the neighbours into account (). In simple terms, nodal eigenvector centrality is decided based on the principal eigenvector, which explains most variance in data. The main principle is that links from important nodes (as measured by weighted degree centrality) are worth more than links from unimportant nodes. All nodes start off equal, but as the computation progresses, nodes with more links start gaining importance. Their importance also influences the nodes to which they are connected. After re-computing many times, the values stabilize, resulting in the final values for eigenvector centrality. Eigenvector centrality was calculated according to the equation below, where *λ* denotes the largest eigenvalue and *e*(*i*) denotes the corresponding principal eigenvector, that is a centrality score *e*(*j*) for each node *i* in an undirected network.

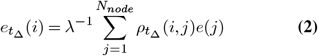

From the above equation, the eigenvector centrality *e*(*i*) of a node *i* is given by the sum of the values within the principal eigenvector corresponding to direct neighbours proportional to the sum of the scores of the node’s network neighbours, where *λ* is a constant and the sum is over all nodes. Eigen-vector centrality is scaled by the proportionality factor *λ*^−1^. Here 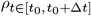 is an element of the correlation matrix representing the strength of the pair-wise correlation between pairs of nodes (*i, j*) within a sliding window. Defining a vector 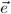 whose elements are the *e*(*i*), results in 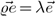, meaning that the vector of centralities is an eigenvector of the correlation matrix.

### B. Genetic Data

Microarray gene expression data from post-mortem healthy human brains were downloaded from the Allan Human Brain Atlas (AHBA) (35). The atlas consist of 926 brain regions; each region tested using an array of 58,692 probes that correspond to 29,181 distinct genes. Among all genes and regions we analysed those of interest for this study. The 926 regions were downsampled to 121 regions of the JBA, using the detailed anatomical labels provided by AHBA. Genes of interest were selected from [The Human Protein Atlas (HPA)](https://www.proteinatlas.org/) database, which generates integrated human gene-associations from curated databases and text mining. The Human Protein Atlas consists of ten separate sections, each focusing on a particular aspect of the genome-wide analysis of the human proteins. We selected for further analysis 71 (out of 84 found in the database) genes associated with Alzheimer’s disease (ADG), which were also mapped by AHBA. Using this procedure we also obtained 13 (out of 17 from the database) genetic variants associated with the cholinergic system AChG. Given the involvement of the cholinergic system in the early stages of AD, we sought to address the question: How do genetic variants associated with the brain cholinergic system contribute to functional network involvement in AD? It should be noted that, the difference between the number of genes found in the HPA and genes used in the analysis comes from the fact that not all genes identified in the database were found in the AHBA atlas.

Using this procedure, we obtained either 121 × 71 or 121 × 13 gene-variants’ matrix, for ADG and AChG variants respectively. These matrices were used for further analysis. To obtain a single vector that explains the most variations in genetic data across brain regions, we performed a principal component analysis (PCA) on the ADG and AChG matrices. We then used the first principal component, for further detailed analysis and the comparisons with the network metrics derived from the fMRI data. To this end, we correlated the first principal component to the node strength and eigenvector centrality cortical maps. In a final step, we selected genes whose expression correlated to the network metrics and visualised common and distinct trends across the measures. Specifically, we constructed *N*_*nodes*_ × *Ngenes/properties* matrix [Fig. 2(A: left panel)].

**Fig. 1.**
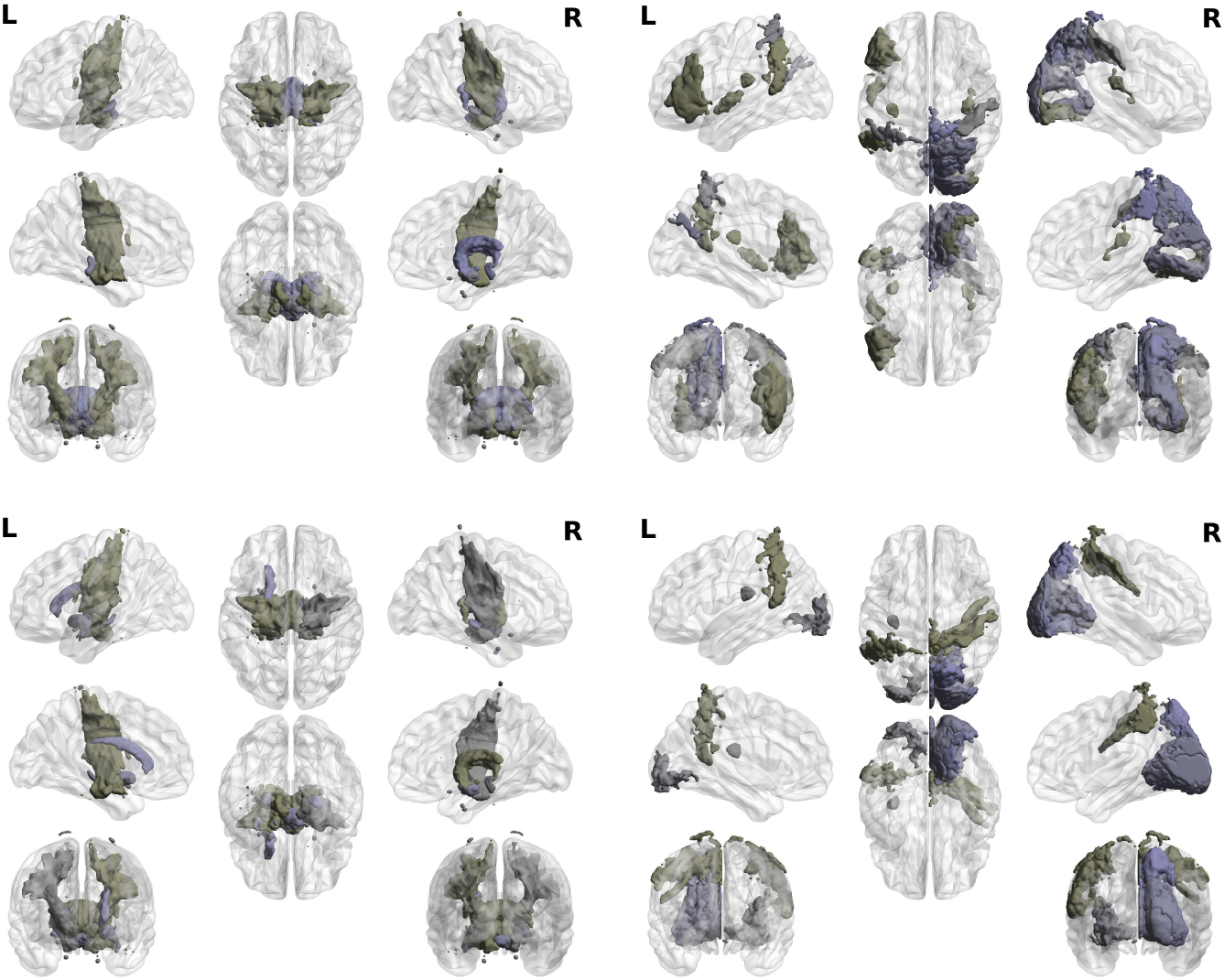
Cortical maps of gene expression across the Juelich Atlas regions. (upper panel) The first principal component of the ACh gene co-variations with the JBA regions. (lower panel) The first principal component of the AD gene co-variations with the JBA regions. In both panels cortical maps of positive coefficients of the PCA are given on the left side and negative on the right. List of regions and their ranks is summarised in Table 2. Common regions for positive coefficients: amygdala group, hippocampal-amygdaloid transition area, corticospinal tract, fornix, lateral and medial geniculate and mamillary body and insular cortex. Differential regions: uncinate fascicle and superior occipito-frontal fascicle (both found only in AD-associated-gene maps).

**Fig. 2.**
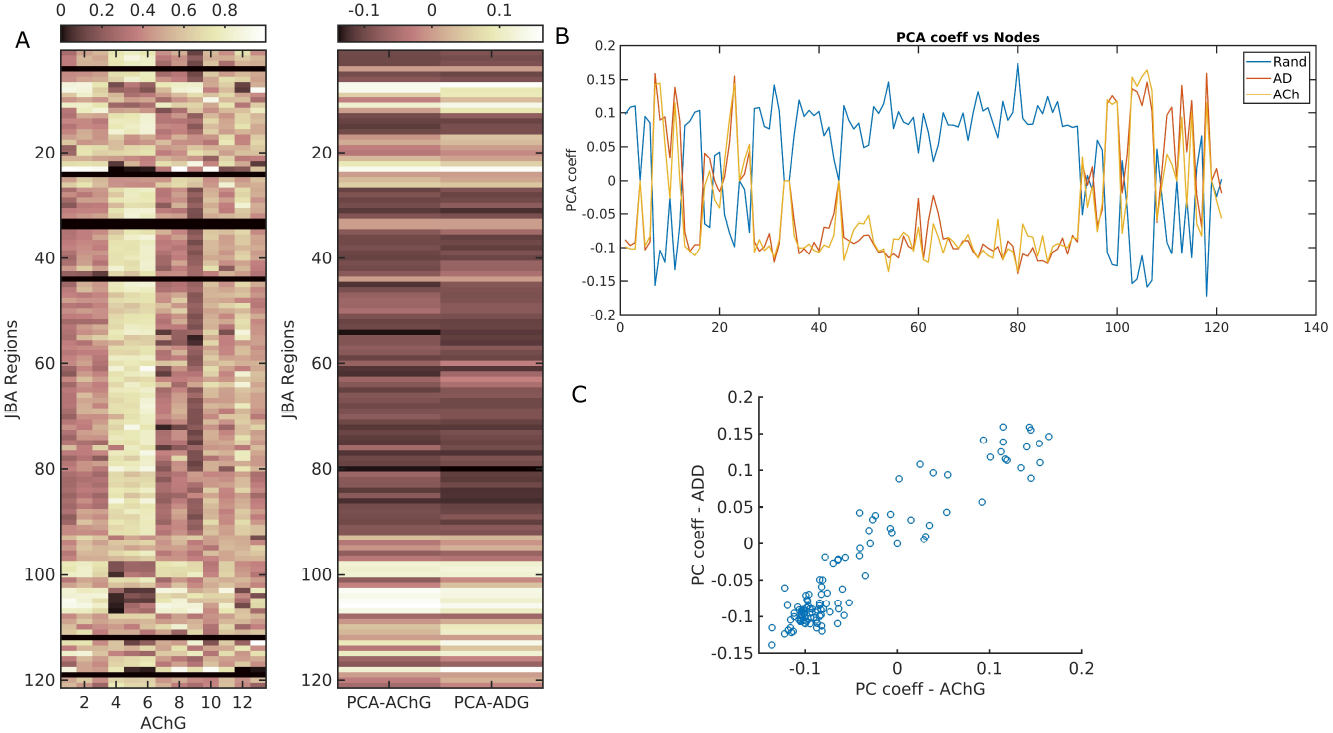
Principal component analysis of gene co-expression across the Juelich Brain Atlas (JBA). (A) Right panel: Heat maps of the first principal component of the AChG and ADG gene co-variations within the JBA regions. List of regions and their ranks is given in SI Table 2. Left panel: Heat map of association between all 13 ACh genes and JBA regions. (B) Plot of the first principal component of the AChG and ADG gene co-variations within the JBA regions. (C) A scatter plot of the correlation between 1^st^ principal component of AChG and ADG polygenic scores for cortical associations. *ρ*=0.95 p << 10^*−*6^.

### C. Statistical Analysis

Our functional networks were constructed on Pearson’s correlation (*ρ*), which represents strength of correlation between regional BOLD signals (time series of regional activity). Due to non-uniform distribution of functional correlations in the brain, network models like ours also show non-normal distribution. Hence, we used non-parametric statistical test to differentiate between nodal measures across individual networks. Changes at the nodal level across cohorts were analysed by means of the non-parametric Kruskal-Wallis (KW) test (36). In addition, to carry this test, we created 100 independent cohort realisations, where each realisation randomly selects 50 participants (with replacement and using a 35 : 15 male-to-female ratio) of each cohort (AD, MCI, and HC). Thus, for each realisation and pair of cortical regions (*i, j*), there are 50 + 50 + 50 = 150 sampled correlations 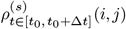 (with *s* = 1,…, 150) representing the (*i, j*)-th functional-link’s weight distribution across the cohorts. The KW null hypothesis is that this data come from the same continuous distribution – i.e., *p*_*KW*_ (*i, j*) > 0.05 – whereas the alternative is that it comes from multiple distributions – i.e., *p*_*KW*_ (*i, j*) ≤ 0.05. As a result, we have a *p*_*KW*_ (*i, j*) matrix for the static correlations 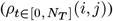 for each realisation (sampled participants) and a set of *p*_*KW*_ (*i, j*; *t*_0_, Δ*t*) matrices for the dynamic correlations 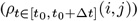.

**Table 2.**
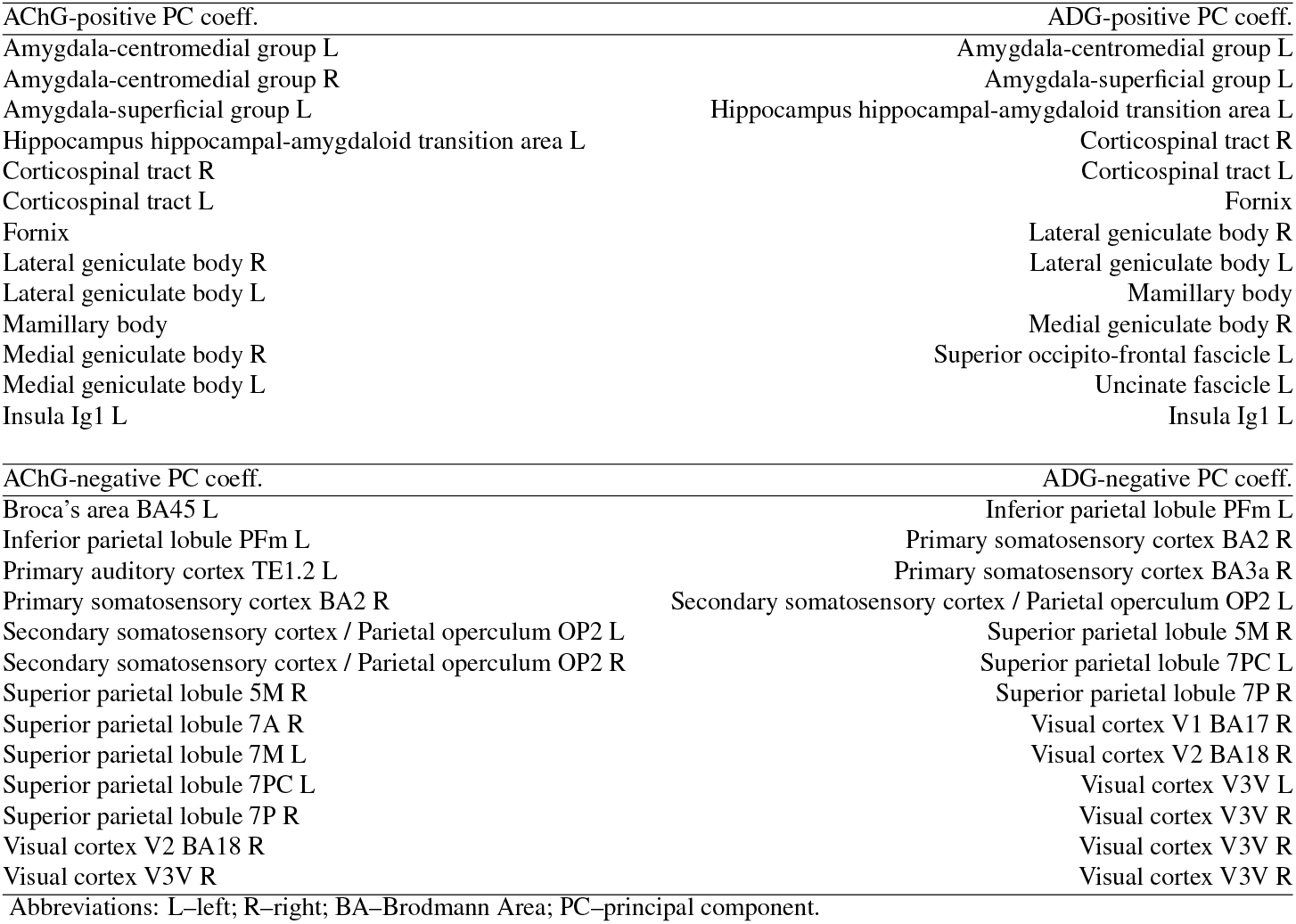
Regions of the Juelich Atlas showing either strong positive or negative associations with gene expression of the cholinergic pathways (AChg) in the brain or Alzheimer’s disease (ADD) A percentage of first 10% regions is shown.

| AChG-positive PC coeff. | ADG-positive PC coeff. |
| --- | --- |
| Amygdala-centromedial group L | Amygdala-centromedial group L |
| Amygdala-centromedial group R | Amygdala-superficial group L |
| Amygdala-superficial group L | Hippocampus hippocampal-amygdaloid transition area L |
| Hippocampus hippocampal-amygdaloid transition area L | Corticospinal tract R |
| Corticospinal tract R | Corticospinal tract L |
| Corticospinal tract L | Fornix |
| Fornix | Lateral geniculate body R |
| Lateral geniculate body R | Lateral geniculate body L |
| Lateral geniculate body L | Mamillary body |
| Mamillary body | Medial geniculate body R |
| Medial geniculate body R | Superior occipito-frontal fascicle L |
| Medial geniculate body L | Uncinate fascicle L |
| Insula Ig1 L | Insula Ig1 L |
| AChG-negative PC coeff. | ADG-negative PC coeff. |
| Broca's area BA45 L | Inferior parietal lobule PFm L |
| Inferior parietal lobule PFm L | Primary somatosensory cortex BA2 R |
| Primary auditory cortex TE1.2 L | Primary somatosensory cortex BA3a R |
| Primary somatosensory cortex BA2 R | Secondary somatosensory cortex / Parietal operculum OP2 L |
| Secondary somatosensory cortex / Parietal operculum OP2 L | Superior parietal lobule 5M R |
| Secondary somatosensory cortex / Parietal operculum OP2 R | Superior parietal lobule 7PC L |
| Superior parietal lobule 5M R | Superior parietal lobule 7P R |
| Superior parietal lobule 7A R | Visual cortex V1 BA17 R |
| Superior parietal lobule 7M L | Visual cortex V2 BA18 R |
| Superior parietal lobule 7PC L | Visual cortex V3V L |
| Superior parietal lobule 7P R | Visual cortex V3V R |
| Visual cortex V2 BA18 R | Visual cortex V3V R |
| Visual cortex V3V R | Visual cortex V3V R |
| Abbreviations: L–left; R–right; BA–Brodmann Area; PC–principal component. |  |

## Results

Our main analyses were based on fMRI data from a sample of 315 individuals from three clinical groups: healthy controls, mild cognitive impairment and Alzheimer’s disease. Dynamic functional networks were analysed across the three groups using a sliding window approach applied to the Juelich Brain Atlas regional BOLD-signal time series. The replication analysis was performed on the same subjects using another brain parcellation, with a lower spatial resolution. Genetic variants associated with Alzheimer’s disease and the cholinergic brain pathways were mapped onto brain regions (of 2 brain atlases), and correlated with the dynamic functional brain networks’ metrics to highlight different measures of cortical organization across different modalities and scales.

### D. Gene Expression

To test the hypothesis that the expression of the genes of interests in the brain is associated with the observed differences in dFC between the three groups, we utilised gene expression maps from the Allan Human Brain Atlas. We used two sets of gene co-expression maps in the brain: one that consists of genes implicated with Alzheimer’s disease (71 genes) and one that consists of gene variants associated with the cholinergic brain pathways (13 genes). To reduce the dimensionality of the genetic data we used their respective principal components, which capture the overall association patterns of the genes × brain regions maps. Fig. 1 shows brain maps of the first principal component coefficients for ADG and AChG co-expression in JBA regions. Here, for the purpose of brevity we show only the first 10 nodes (with the highest coefficient) for both maps. Although, visually very similar, the two maps differ in the regions mapped out. Interestingly, most of the regions are spatially very close to one another. Common regions for positive coefficients: amygdala group, hippocampalamygdaloid transition area, corticospinal tract, fornix, lateral and medial geniculate and mamillary body and insular cortex. Differential regions: uncinate fascicle and superior occipito-frontal fascicle (both found only in AD-associated-gene maps).

Based on 71 genes associated with AD, we found that the first principal component of gene expression explains 51.16% of co-expression variance. Adding of further four components explained 72.20% of total variance. Interestingly, 13 genetic variations associated with the cholinergic system in the brain explained 72.33% of co-variations across the JBA cortical parcellation (where the first five components explained 92.06% of the total variance in data). Figure 2 depicts heatmaps of AChG × JBA regions co-variations and coefficients of the first principal component of AChG and ADG associations. Their correlations with the dFC measures are described section “Association between Functional Networks and Gene Expression”.

The first principal component coefficients of ADG and AChG patterns of variations associated with H-OBA cortical regions is shown in Supplementary Fig. 5. Given that only sub-cortical H-OBA regions’ labelling is lateralised, separate analysis was performed on either sub-cortical or cortical regions of this atlas. The patterns of associations, in terms of their left-right asymmetry are very similar to those obtained for the JBA (see Fig. 1). It should be also noted that the separate mapping of cortical and sub-cortical regions of the H-OBA showed that AChG are predominately associated with more central regions, such as parahippocampus and cingulate cortices, and ADG are predominately associated with the cortical surface regions of the, e.g, occipital and temporal lobes (see Supplemant Table 4).

### E. Association between Functional Networks and Gene Expression

In addition to the brain-gene-expression maps, we also examined the relationship between maps and dFC metrics of interest. In particular, building on the emerging gene-expression and cortical-architecture relations, we examined association between the dFC metrics (the node strength and eigenvector centrality) and the first principal component of the ADG and AChG cortical variations. This was done by correlating the first principal components of genetic × regional co-variations with the pair-wise contrast between either the node strength or eigenvector centrality across the three groups, providing in total of 6 values for each measure indicating weights of correlations. Table 3 shows correlations between the first principal component and the two network measures, for the polygenic risk (ADG) and for the cholinergic pathway genes (AChG). As an illustration, Figure 3, shows the correlation between AD/HC contrast for eigenvector centrality and AChG and ADG principal components. Given the high similarity in the two graphs we also calculated correlation between the two principal components, which reached a value of 0.95 (p << 106). We focused only on the first principal component, because in both cases they explain the highest portion of the variance in gene expression in the brain. We argue that the relatively high AChG co-expression in the brain (quantified by the 1st PC) most likely reflects a property of the JBA parcellation, which includes the superficial white matter fibers just beneath the specific cytoarchitecturally defined cortical areas. Another, equally possible explanation is the division of the hippocampus into 10 sub-regions, where the 1st PC of AChG co-expression across the JBA, could map cholinergic signaling in the hippocampus (see for example (37)).

**Table 3.**
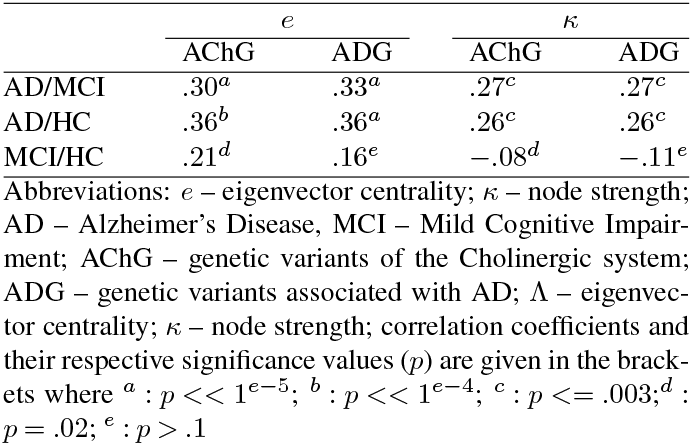
Correlation analysis for the first principal component (z-score) of gene expression data and the two dFC metrics, eigenvector centrality (z-score) and node strength (z-score).

| | $e$ | | $\kappa$ | |
| --- | --- | --- | --- | --- |
|  | AChG | ADG | AChG | ADG |
| AD/MCI | .30<sup>a</sup> | .33<sup>a</sup> | .27<sup>c</sup> | .27<sup>c</sup> |
| AD/HC | .36<sup>b</sup> | .36<sup>a</sup> | .26<sup>c</sup> | .26<sup>c</sup> |
| MCI/HC | .21<sup>d</sup> | .16<sup>e</sup> | -.08<sup>d</sup> | -.11<sup>e</sup> |

Abbreviations: $e$ – eigenvector centrality; $\kappa$ – node strength; AD – Alzheimer’s Disease, MCI – Mild Cognitive Impairment; AChG – genetic variants of the Cholinergic system; ADG – genetic variants associated with AD; $\Lambda$ – eigenvector centrality; $\kappa$ – node strength; correlation coefficients and their respective significance values ( $p$ ) are given in the brackets where <sup>a</sup> : $p < 1e^{-5}$ ; <sup>b</sup> : $p < 1e^{-4}$ ; <sup>c</sup> : $p \leq .003$ ; <sup>d</sup> : $p = .02$ ; <sup>e</sup> : $p > .1$

**Fig. 3.**
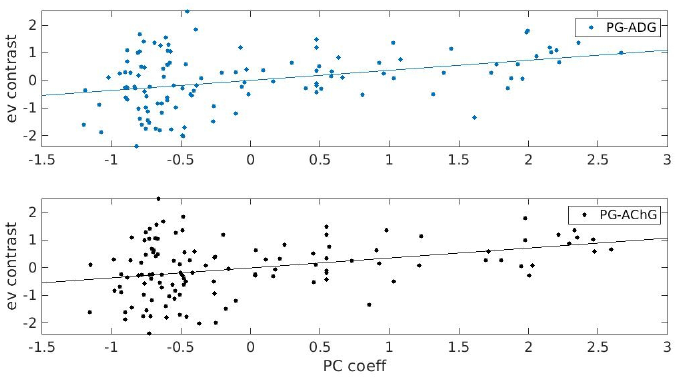
The correlation between dFC eigenvector centrality and genetic variations. Contrast in eigenvector centrality (z-score) between AD and HC group was correlated with the first principal component (z-score) of gene expression associated with AD (upper panel) and cholinergic system (lower panel).

### F. Dynamic Functional Networks by Clinical Groups

Finally, we briefly describe behaviour of the dFC networks in the three groups using the two centrality measures: the node strength and eigenvector centrality. Fig. 4 shows the 2 network metrics across 121 regions (nodes) of the JBA, when averaged across the sliding windows. We observed statistical differences between the groups at the local (nodal) level: The dFC node strength across the white matter JBA regions differ between the AD and MCI and AD and HC groups, where the AD subjects show higher values than the other two groups. Similar statistical differences also exists for the JBA visual cortex areas, but the AD subjects showed lower node strength than the other two groups. The eigenvector centrality revealed more ‘irregular’ differences, with the parietal and the occipital nodes’ being consistently different when comparing AD and MCI and HC subjects. A more comprehensive analysis of the dFC in the three groups (38), showed how the fluctuations in the dFC pair-wise links evolve over the acquisition time. Here, we only show the averaged values of these measures for the purpose of visualising their values across the JBA regions.

**Fig. 4.**
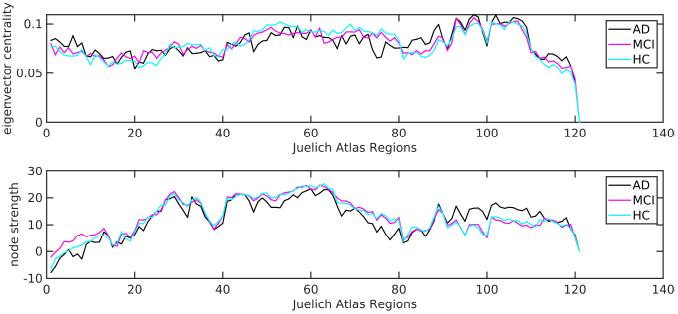
Dynamic functional connectivity network metrics in in AD, MCI and HC groups. (Upper panel) eigenvector centrality and (lower panel) node strength, shown when averaged over the 121 Juelich Brain Atlas regions.

**Fig. 5.**
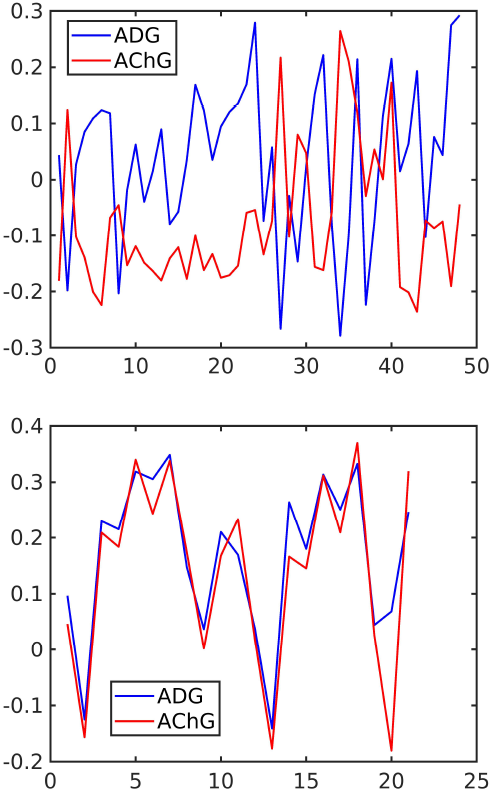
First principal components of gene expression association with (upper panel) 48 cortical regions of the Harvard-Oxford Atlas and (lower panel) 21 subcortical regions of the H-O brain atlas. Abbreviations: ADG – AD-associated genes; AChG – Cholinergic-pathways genes

## Discussion

Here, we examined putative associations between genetic variations associated with Alzheimer’s disease and dynamic functional connectivity from data of 315 individuals from three age-matched and sex-balanced clinical groups: AD, MCI, and healthy seniors. Our results provide evidence of the relationship between dynamic functional connectivity and the gene expression in the brain associated with AD, especially those co-expressed in the cholinergic pathways. We show that measures of centrality of any given node across the dynamic functional connections, correlate with the level of gene expression. Higher gene expression is associated with higher positive contrast in nodal importance between AD and MCI and HC, and this was most prominent in white matter regions. Lower genetic risk is associated with the higher (by absolute value) negative contrast between the AD and MCI/HC groups in the parietal and visual areas.

Dynamic functional connectivity has been identified as a better predictor of cognitive impairment in AD patients, compared to conventional static FC analysis. Numerous studies have provided compelling evidence that the classification of AD from MCI or HC subjects significantly improved when using the dFC networks as a defining feature of altered brain activity in AD (5, 39–41). However, despite this evidence for its utility, how dFC relate to the brain biology at the microscale remains elusive. In this study, we investigated dFC network metrics and genetic variations associated with AD, in an attempt to identify how large-scale network dynamic in AD relates to the microscale properties of cortical vulnerability to the disease, i.e., to variations in the regional gene expression. Our aim was to add a new, genetic dimension to the differences in dFC, which has been established between AD patients and healthy subjects, yet only in terms of dynamic metastability and nodal importance in the dFC. We aimed specifically to compare, dynamic metastability and nodal importance using the parcellation of the Juelich cytoarchitectonic atlas. This is a multimodal brain atlas created to delineate white matter fibres’ pathways with known associated functions, as well as major white matter tracts and grey matter regions (32).

An important question in mapping neurodegenerative processes in AD remains — how the brain maps of gene expression are related to brain functional connectivity maps. By incorporating information about regional gene expression in the brain to annotate dFC changes in AD and MCI subjects, neural gene expression maps have been created as innovative tools explaining sources of variations in neuroimaging features across a range of neurodegenerative disorders, including Parkinsonism and dementia (42–44). Here, we provide the first attempt to explore this issue mapping genes and functional connectivity in the AD continuum. For this, we incorporated information about regional gene expression in the brain to annotate dFC changes and contrasted these with MCI and control subjects. First, we produced gene expression maps onto the JBA regions, using genetic variants associated with AD (71 genes in total). Secondly, we also mapped genes associated with the cholinegic brain pathways (13 genes). This is based on the rationale that during MCI and AD there is a progressive loss of forebrain cholinergic neurons giving rise to a widespread cortical dysfunction (for review see (45, 46)). Interestingly, the regions showing AD-related-polygenic association largely overlap with those showing cholinergic-pathways-related genetic association and include: amygdala group, hippocampal-amygdaloid transition area, corticospinal tract, fornix, lateral and medial geniculate and mamillary body and insular cortex. That is, the maps are similar at the regional level, but they usually show differences in the regional sub-areas and/or their interhemispheric homologous. While regions associated with the cholinergic pathways gene co-expression are highly symmetrical and found in both hemisphere equally, regions associated with AD-related genetic variations are asymmetrically distributed across the hemispheres (see Table 2). Areas with highly emotional functions are more associated with AD genes in the left hemisphere, sensory information processing is more in the right. This is the first study of this kind to link cholinergic genetic associations with fMRI. The data therefore suggest that there is no hemisphere-selective vulnerability to cholinergic degeneration and functional loss, but rather both hemispheres are affected in the same manner in AD. Our findings are consistent with early reports on the extent of left-right symmetry in the cholinergic deficits in AD brains, which was found to be symmetrically distributed compared to more asymmetrical morphological lesions (47, 48). Table 2 shows that it is the ADG variants which predominantly map out regions in the left hemisphere. In patients with AD, the left hemisphere is more severely affected, both structurally and metabolically (49–51). In addition, a trend for faster grey matter loss in the left hemisphere was also observed in age matched controls (52). Our results indicate that the variations in lateralisation of the brain tissue loss in healthy ageing, but also in AD, may be explained by AChG and ADG variants expression across the brain structures. As supported by early studies in AD, such loss is more symmetrical at the early stages, while becoming predominantly associated with the left hemisphere as the disease progresses (20). We have previously reported similar heterogeneity in brain atrophy across the cortical surface, albeit in different from of dementia (16) as well as in healthy ageing (53). This was validated across the H-OBA sub-cortical regions (see Table 4), where seven out of ten regions associated with ADG variants are in the left hemisphere. However, given the differences in the resolutions, but also in labelling the regions in the two atlases, these results should not be considered as 1:1 mapping at the regional level.

**Table 4.** Sub-cortical and cortical regions of the Harvard-Oxford Brain Atlas which show strong positive associations with gene expression of the cholinergic pathways (AChG) in the brain or Alzheimer’s disease (ADG). The first 10 regions are listed for both divisions.

| ACh-positive PC coeff. | ADG-positive PC coeff. |
| --- | --- |
| sub-cortical hubs |  |
| left pallidum | right pallidum |
| right pallidum | left caudate |
| left caudate | left pallidum |
| right caudate | right accumbens |
| left putamen | right caudate |
| right lateral ventricle | left putamen |
| right putamen | left accumbens |
| right accumbens | right putamen |
| left lateral ventricle | left lateral ventricle |
| left thalamus | left thalamus |
| brain-stem | left amygdala |
| cortical hubs |  |
| parahippocampal gyrus, anterior | occipital pole |
| subcallosal | intracalcarine |
| parahippocampal gyrus, posterior | supracalcarine |
| occipital fusiform gyrus | cuneal |
| insular | occipital fusiform gyrus |
| lingual gyrus | lingual gyrus |
| cingulate gyrus, anterior | parietal operculum |
| temporal fusiform, posterior | lateral occipital, inferior |
| cingulate gyrus, posterior | postcentral gyrus |
| temporal occipital fusiform | precuneus |
| temporal fusiform, anterior | lateral occipital, superior |

**Table 5.**
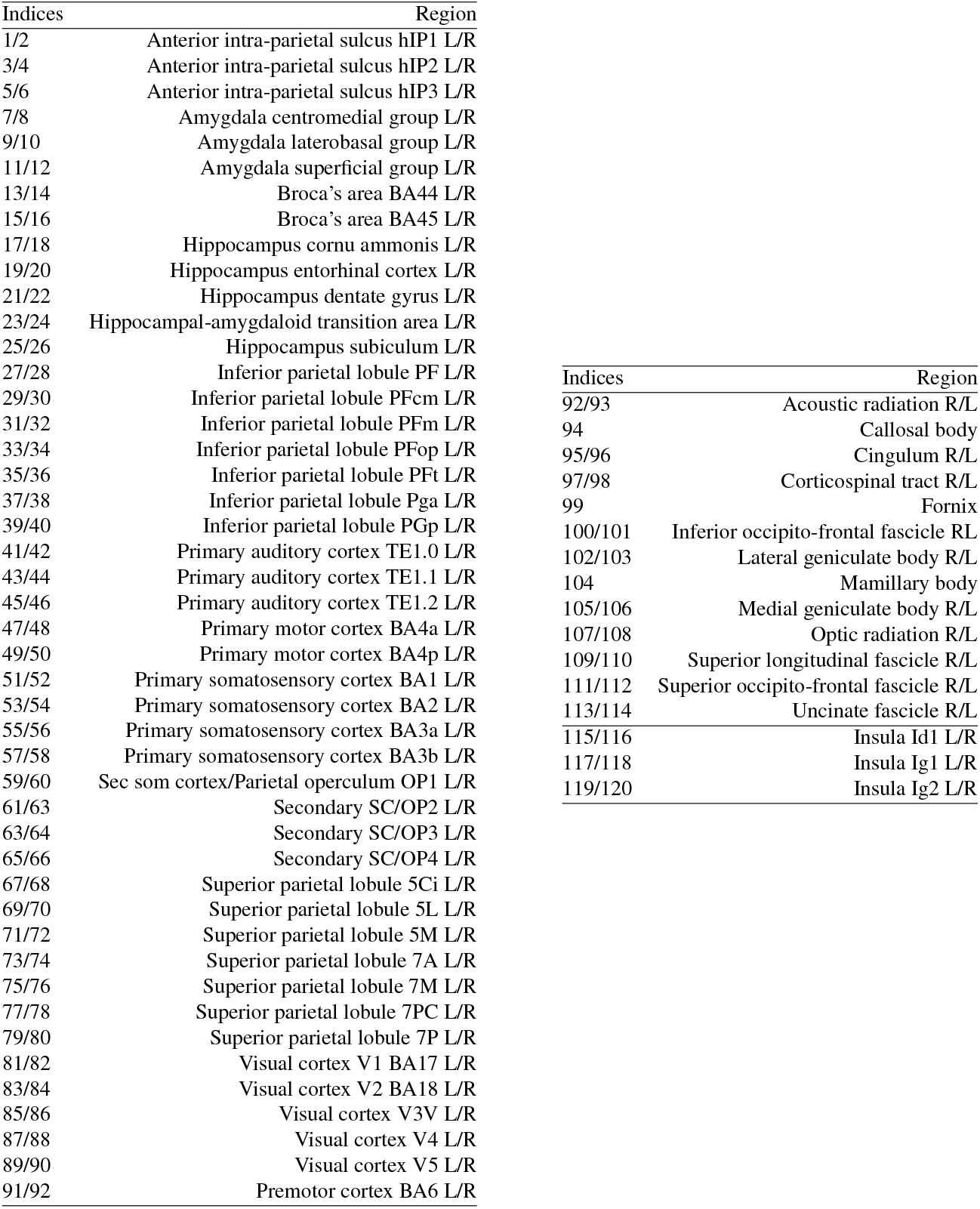
Juelich Brain Atlas regions. The mid lines indicate border between grey and white matter regions.

## Conclusion

In summary, we aimed to investigate the relationship between genetic variations and dynamic functional connectivity in Alzheimer’s disease, mild cognitive impairment and healthy individuals. Linking gene expression and cortical regions provided evidence of variations in terms of hemisphere-selected vulnerability to AD. Furthermore, our findings indicated similarity in genetic variations associated with Alzheimer’s disease and also associated with the brain cholinergic pathways and the drivers of dynamic functional connectivity aberrations in the disease. The results may provide a possible genetic substrate for changes in the metastable dynamic of cortical networks in AD, and provide evidence of genetic associations with asymmetric changes in the cortical surface in healthy ageing and Alzheimer’s disease. Our work uncovers a fundamental feature of the brain connectivity organization at differential scales.

## Data Availability

All data produced in the present study are available upon reasonable request to the authors

## Competing interests

No competing interest is declared.

## Author contributions statement

V.V. conceptualised the study, analysed genetic and fMRI data, drafted the manuscript and supervised the project. N.R. conducted analysis of functional MRI data. V.V and G.R. wrote and reviewed the manuscript.

## Supplementary Note 1: Supplementary Material

### A. Gene expression mapped onto the Harvard-Oxford Atlas regions

Validation of gene expression in the JBA regions, was performed using the Harvard-Oxford Atlas (H-OBA), one of the atlases implemented in FSL that is based on similar probability maps but with the lower resolution. The H-OBA is a probabilistic atlas covering 48 cortical and 21 subcortical structural areas, derived from structural data and segmentation, provided by the Harvard Center for Morphometric Analysis (see for example (54)). Given that the labelling of the H-OBA, which does not differentiate left and right cortical regions, but only sub-cortical, we performed analysis on either 48 cortical or 21 sub-cortical regions separately. This was done using the same approach as described in Section B.

We used two different polygenic data sets, whose variations were mapped onto the brain: one that consists of genes implicated in Alzheimer’s disease (71 genes in total) and the oter one that consists of gene variants implicated in the cholinergic brain pathways (13 genes in total). To reduce the dimensionality of the genetic data, we used their respective principal components, which capture the overall association patterns genes × brain regions (for 2 brain atlases). Based on 71 genes associated with AD (or ADG in the text), we found that the first principal component explains 41.69% of co-expression variance (while the first three components have explained 66.71% variances). Similarly to the gene expression analysis for JBA regions, 13 genetic variations associated with the cholinergic system in the brain (AChG), explained 50.39% of co-variations across the JBA cortical parcellation (where the first two components explained 19.06% of the total variance in data). In the subsequent analysis of gene expression across 48 cortical and 21 subcortical regions separately, the following patterns emerged: The first principal component of AChG associations with the H-OBA regions explained 68.68% (across 48 cortical regions) and 48.89% (across 21 subcortical regions) of variance. The first principal component of ADG associations with the H-OBA regions explained 41.68% (across 48 cortical regions) and 52.22% (across 21 subcortical regions) of variance.

## Notes

### Competing Interest Statement

The authors have declared no competing interest.

### Funding Statement

This work is supported by funds from Roland Sutton Academic Trust (RG:\#RG13688 and \#DSR1058-100).

### Author Declarations

Source data were openly available before the initiation of the study. http://adni.loni.usc.edu/data-samples/data-types/mri https://www.proteinatlas.org/

